# Understanding COVID-19 spreading through simulation modeling and scenarios comparison: preliminary results

**DOI:** 10.1101/2020.03.30.20047043

**Authors:** Cesar Bordehore, Zaida Herrador, Eva S. Fonfría, Miriam Navarro

**Author notes:** Corresponding author for modeling aspects: Cesar Bordehore, PhD Biology. Corresponding author for medical aspects: Miriam Navarro, MD, PhD.

## Abstract

Since late 2019 the world is facing the rapid spreading of a novel viral disease (SARS-CoV-2) provoked by the coronavirus 2 infection (COVID-19), declared pandemic last 12 March 2020. As of 27 March 2020, there were more than 500,000 confirmed cases and 23,335 deaths worldwide. In those places with a rapid growth in numbers of sick people in need of hospitalization and intensive care, this demand has over-saturate the medical facilities and, in turn, rise the mortality rate.

In the absence of a vaccine, classical epidemiological measures such as testing, quarantine and physical distancing are ways to reduce the growing speed of new infections. Thus, these measures should be a priority for all governments in order to minimize the morbidity and mortality associated to this disease.

System dynamics is widely used in many fields of the biological sciences to study and explain changing systems. The system dynamics approach can help us understand the rapid spread of an infectious disease such as COVID-19 and also generate scenarios to test the effect of different control measures.

The aim of this study is to provide an open model (using STELLA® from Iseesystems) that can be customized to any area/region and by any user, allowing them to evaluate the different behavior of the COVID-19 dynamics under different scenarios. Thus, our intention is not to generate a model to accurately predict the evolution of the disease nor to supplant others more robust -official and non-official-from governments and renowned institutions. We believe that scenarios comparison can be an effective tool to convince the society of the need of a colossal and unprecedented effort to reduce new infections and ultimately, fatalities.

## INTRODUCTION

Since late 2019 the world is facing the rapid spreading of a novel viral disease (COVID-19) provoked by the coronavirus 2 (SARS-CoV-2) infection. On 12 March 2020, this infectious disease was declared pandemic by the World Health Organization (WHO). As of 27 March 2020, there were more than 500,000 confirmed cases and 23,335 deaths worldwide. An updated daily report can be seen at https://www.who.int/emergencies/diseases/novel-coronavirus-2019/situation-reports/.

The case fatality rate of COVID-19 varies depending on the country and the state of the pandemic (updated information at https://www.cebm.net/covid-19/global-covid-19-case-fatality-rates/) and it oscillates generally around 3% and 6% with some exceptions such as Italy, where it reaches 10%. The case fatality rate (CFR) was established at 1.6% in South Korea, 3.04% in China, 10.09% in Italy, and 7.38% in Spain, but the latter might be overestimated since many mild cases have not been spotted. CFR may depend on the quality of the health care received by the patient, detection capacity of each country, previous conditions and age of the patient and also, and probably the most relevant number, the effort in diagnosing, that would rise up the numbers of confirmed infected people (WHO 2020a).

What seems to be clear is that, in those places were there has been a rapid growth in numbers of sick people in need of hospitalization and intensive care, this demand has over-saturated the medical facilities and availability of specialists which in turn, has risen the mortality rate by the shortage of health care resources. This dramatic scenario can be seen now in Italy, where the shortage in ICU (Intensive Care Unit) increased the crude mortality ratio up to a 7.7% (Lazzerrini and Putoto, 2020).

In the absence of a vaccine, classical epidemiological measures such as testing in order to isolate the infected people, quarantine and physical distancing of people are ways to reduce the growing speed of new infections as much as possible and as soon as possible. Thus, this measure is being a priority for all governments in order to minimize the morbidity and mortality associated to this disease (Ferguson et al. 2020, WHO 2020a).

System dynamics is widely used in many fields of the biological sciences to study and explain changing systems, such as population dynamics, predator-prey dynamics, spread of diseases or pests (Hannon and Ruth, 2009). The system dynamics approach can help us understand the rapid spread of an infectious disease such as COVID-19 and also generate scenarios in the model to test the effect of different control measures.

The aim of this study is to provide an open model that can be customized to any area/region and by any user, allowing them to generate scenarios to compare different intervention measures and evaluate the different behavior of the COVID-19 dynamics. Thus, our intention is not to generate a model to accurately predict the evolution of the disease nor to supplant others more robust -official and non-official-from governments and renowned institutions. We believe that scenarios comparison can be an effective tool to convince the society of the need of a colossal and unprecedented effort to reduce new infections and ultimately, fatalities.

## METHODS

### 1. MODEL STRUCTURE AND CUSTOMIZATION

We used data of COVID-19 spread from Spain to calibrate the model and show how to generate scenarios.

To build the model, we used the program STELLA® (IseeSystems) version 9.0.2. This permits to generate a model that can be adapted to all kind of scenarios by anyone with a basic knowledge of STELLA®, since our intention is also to promote participation of other scientists, health care workers and policy-makers to improve the structure and accuracy of the model.

We decided to use a SEIR (Susceptible, Exposed, Infected, Recovered) model adapted to the complexity of the COVID-19 spread.

STELLA® works with 4 main items: Stocks, *Flows, Converters* and *Connectors* (Figure 1):

**Figure 1.**
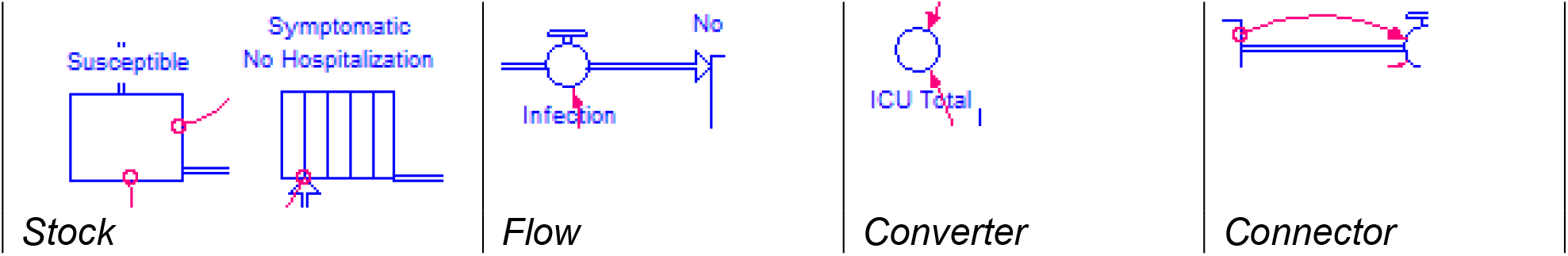
Types of main items used in the STELLA model.

- The *Stocks* represent something that accumulates people in this case (mainly), although they can also refer to materials, money, etc. At any time, the *Stocks* reflect the numbers of items. They are represented by rectangles.
- *Flows* are the inputs and outputs of the *Stocks*. They reflect the number of items that enters or exits a *Stock* per unit of time. In this model we use ‘day’ as a unit of time. They are represented by a wide arrow with a valve.
- *Converters* are used to modify an activity; they are numbers (e.g. a constant) or formulae that will change their result depending on other parameters (i.e. other *Converters* or *Stocks* value). The symbol is a red circle.
- The *Connectors* transmit an input or an output from/to a *Converter, Stock* or *Flow*. They are represented by a red thin line.

Complete tutorials of the program can be downloaded from the official site (www.iseesystems.com) and other concise ones such as the one from Shiflet and Shiflet (2014). There are also useful video tutorials on the internet at www.iseesystems.com and in YouTube (e.g. for a short introduction to system dynamics models in STELLA https://youtu.be/IenySRdkRu8 and a more comprehensive tutorial https://youtu.be/MXSPiR8uSAo). We also recommend the book *Dynamic modeling of Diseases and Pests* (Hannon and Ruth, 2009).

The general structure of the model is shown in Figure 2. The STELLA file can be downloaded from Supplementary Material (Bordehore_et_al_ Understanding_COVID_19_spreading_v29_03_2020).

**Figure 2.**
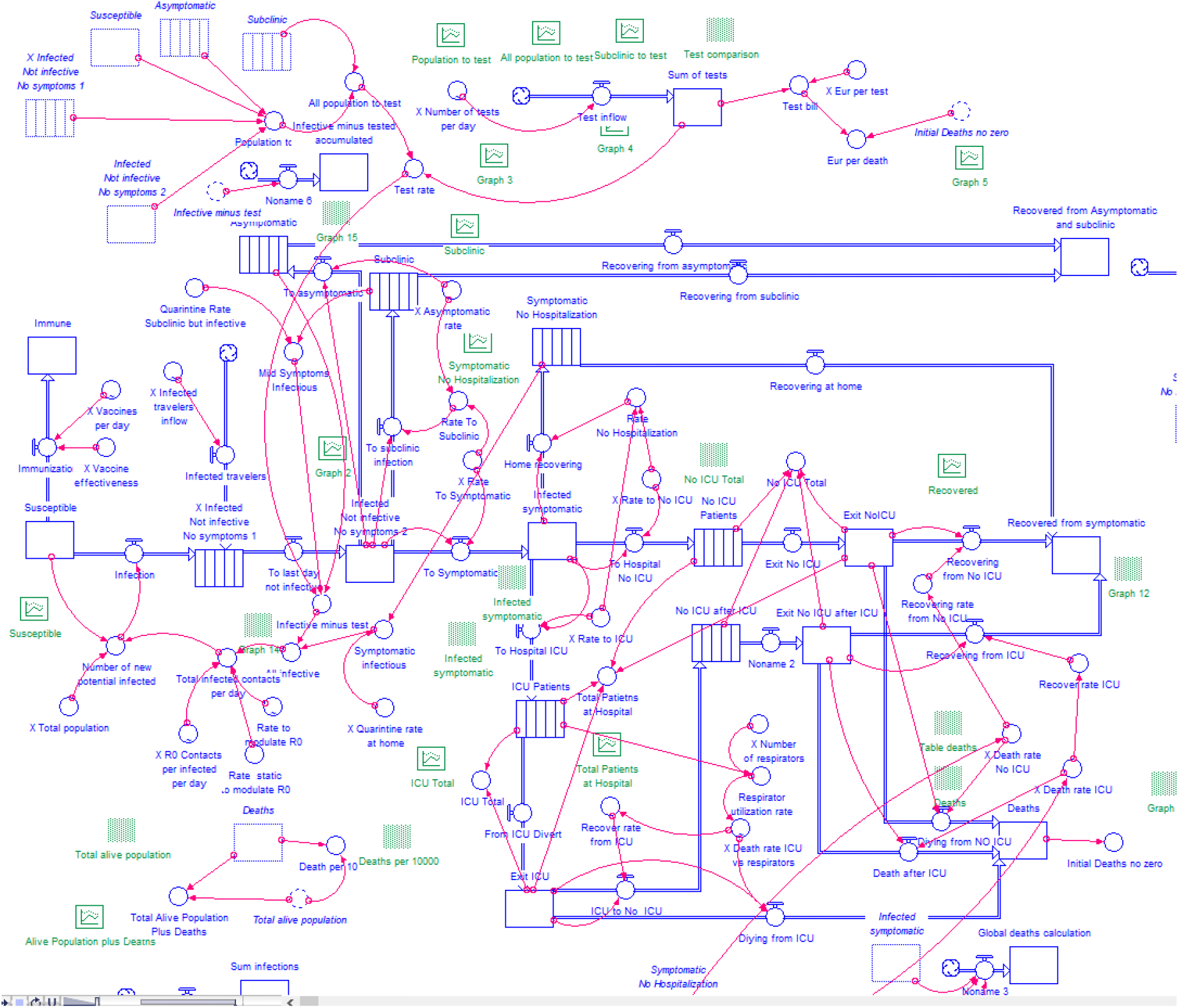
Structure of the STELLA model for COVID-19 spreading.

In Table 1 we describe the *Stocks* used in the model and how we obtained each parameter.

**Table 1.**
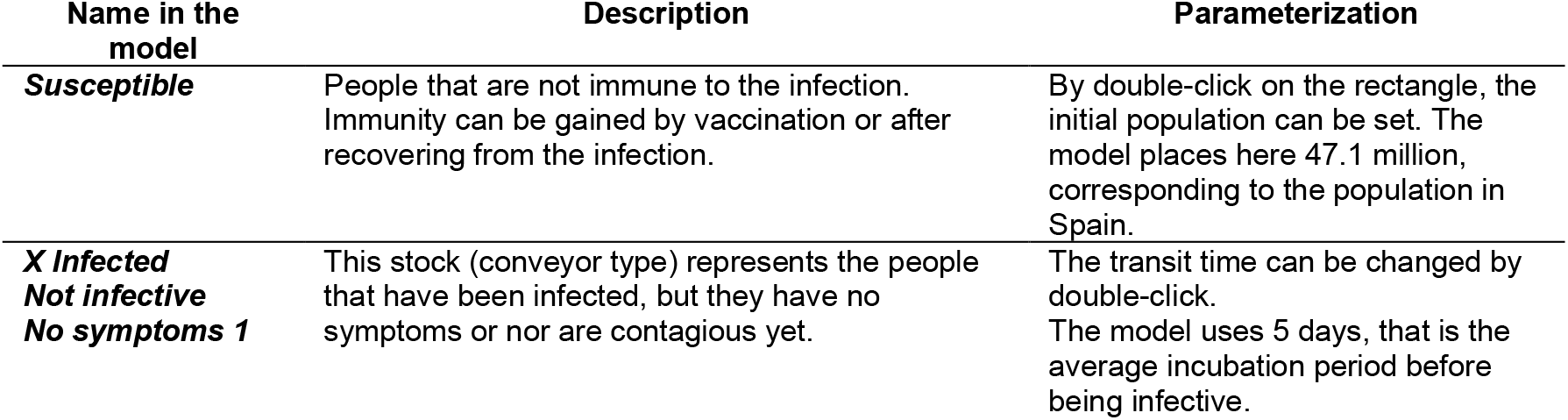
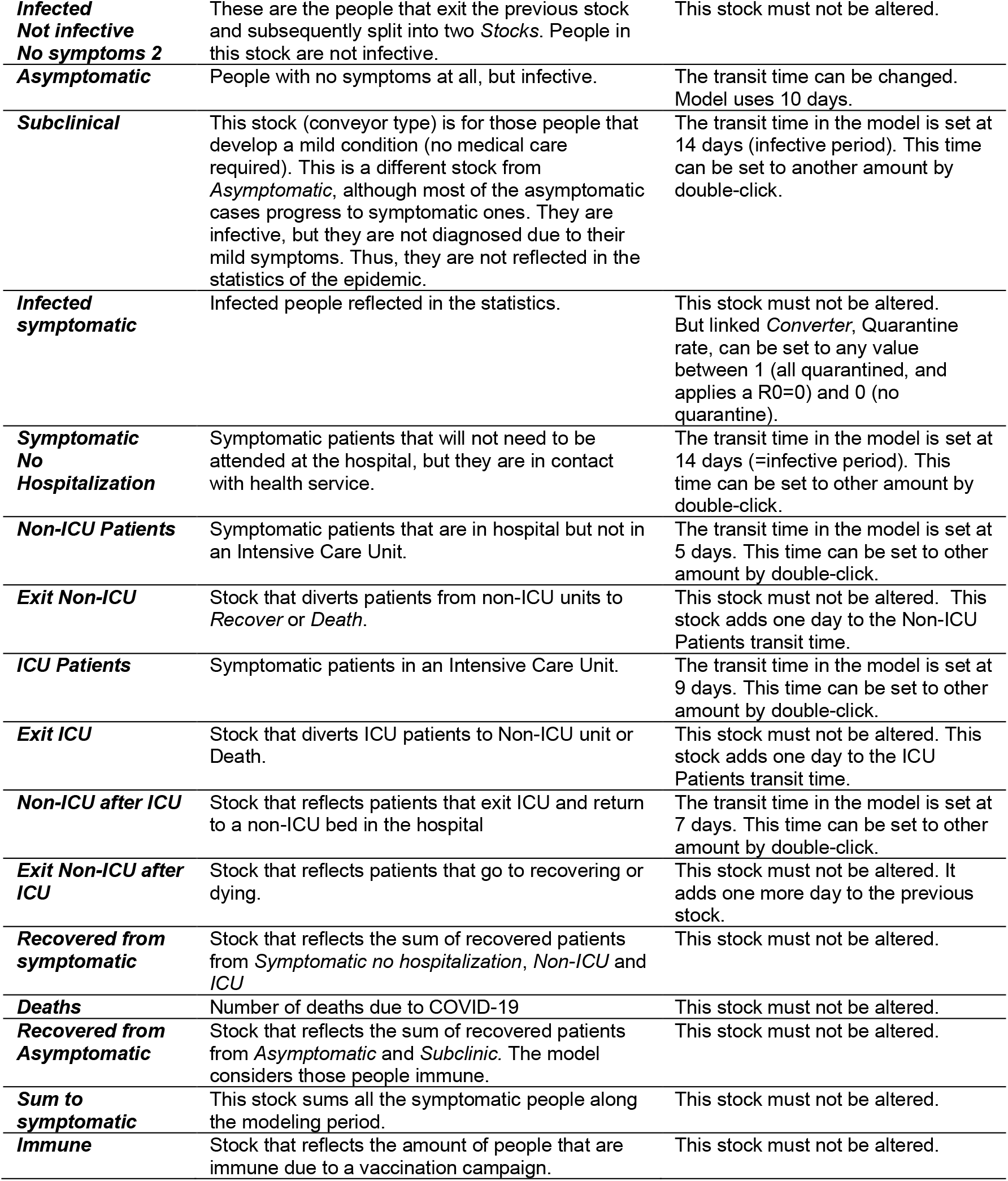
Model *Stocks* description and parameterization.

The model can be customized by just modifying some values of *Converter*s and *Stocks* (name beginning with an X in the model). All the auxiliary variables can be edited by double-click on top of it.

### 2. MODEL CALIBRATION

We used official data from the Spanish Ministry of Health (MSCBS, 2020) to calibrate the model. Model adjustment was performed by using the information provided in Tables 1, 2 and 3.

**Table 2.**
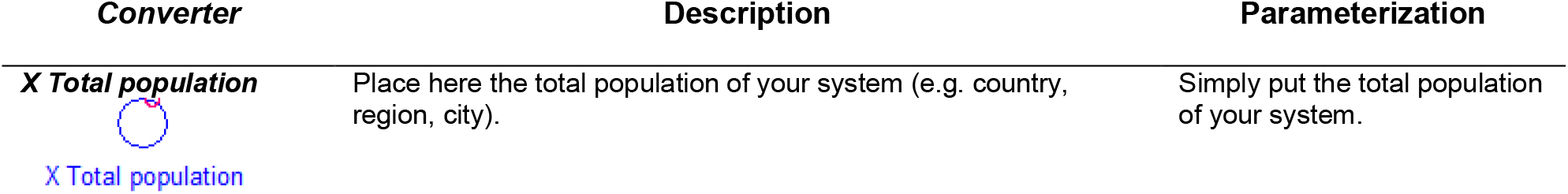
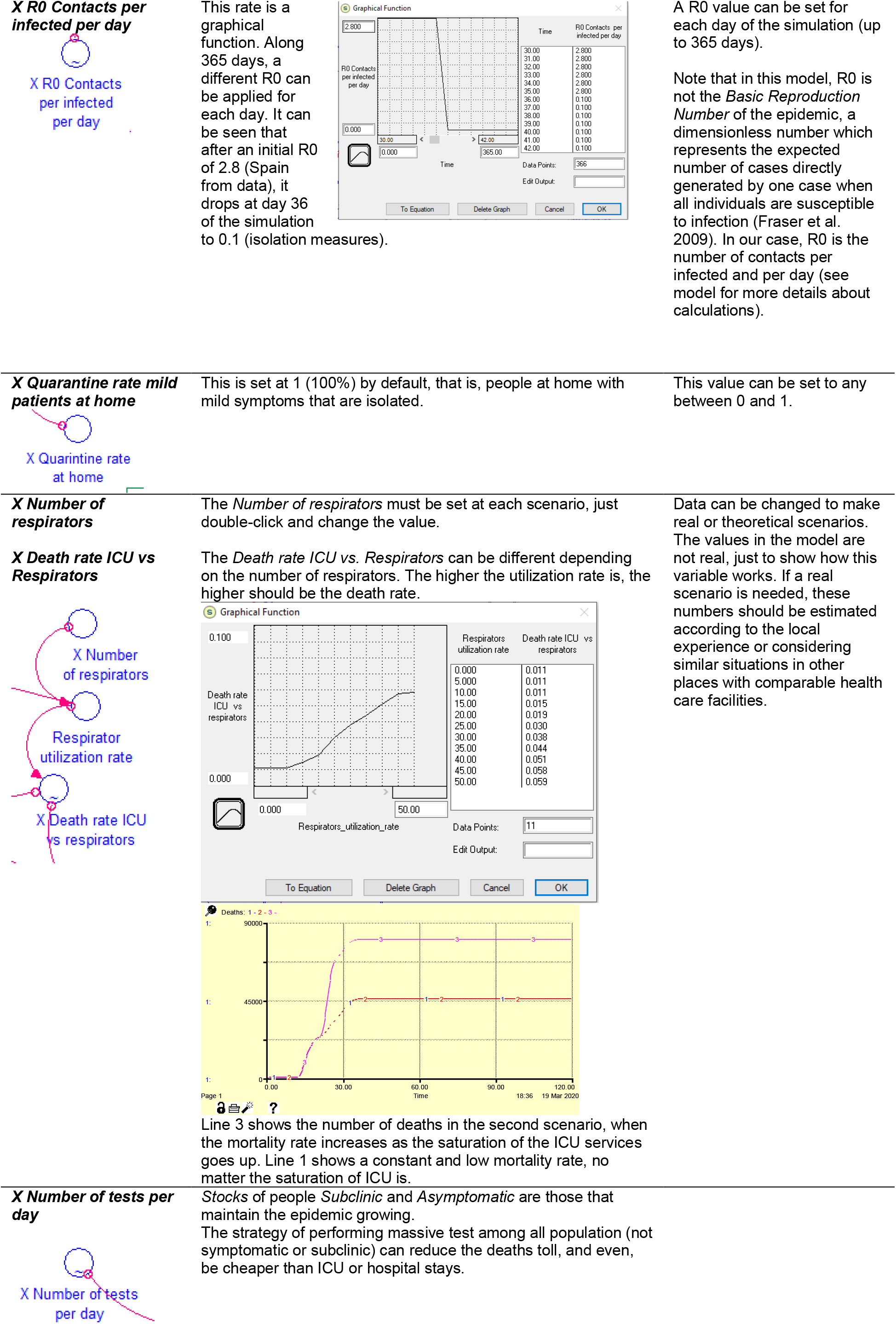
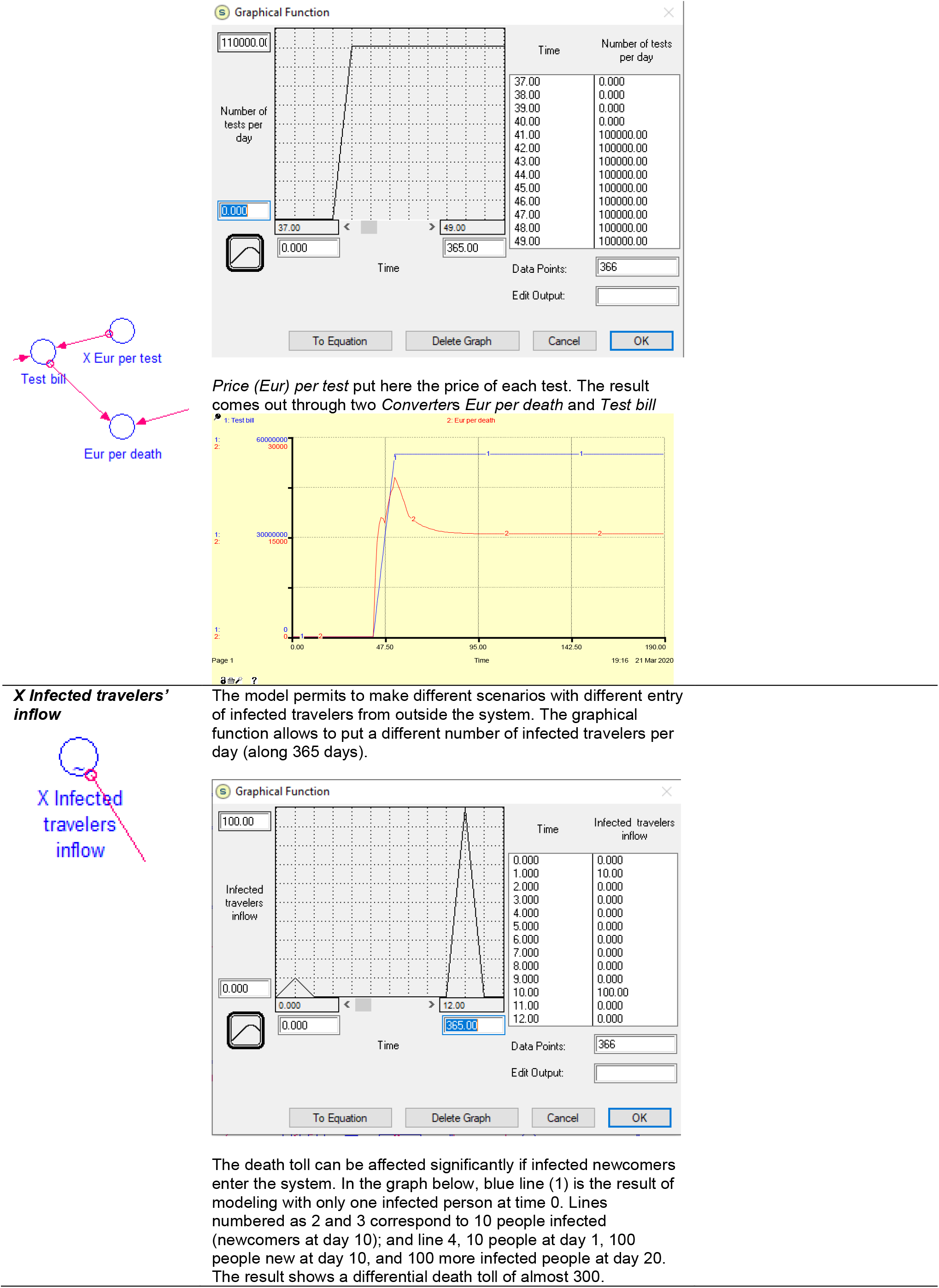
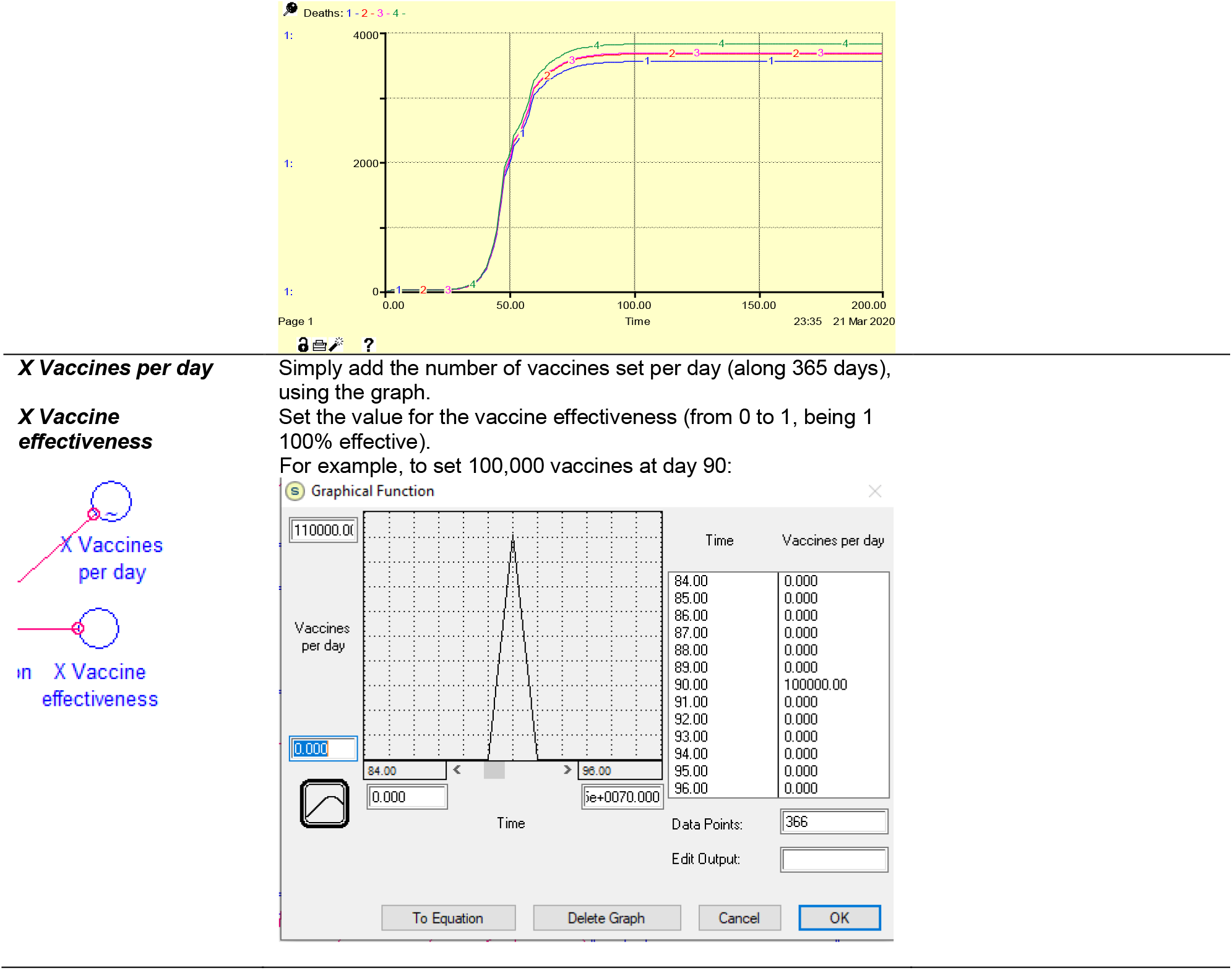
Model main *Converter*s and parameterization.

**Table 3.** Basic parameters to seed the model of the Spanish spread of COVID-19. The customizable parameters are labelled with an “X” at the beginning.

| Parameter | Numbers | Source |
| --- | --- | --- |
| **X Total population** | 47,100,000 | Spanish National Statistics Institute (INE 2020). |
| **X Infected**<br>**Not infective**<br>**No symptoms 1** | 1 | This is the initial number of infected people. This number does not pretend to be the real number of infected people entering Spain, it is just the seed for the geometric progression of the number of infected people. Afterwards, we adapted the time in the progression to the real date according to real numbers. |
| **X R0 Contacts per infected per day** | Graphical function. $R_0=2.33$ before 14/03/2020. | Own estimation, see Figure 3. |
| **X Rate to symptomatic** | 0.626 (=62.6%) | Own estimation calculated from a 3.4% death rate at 21/03/2020 (25,374 cases, 1,378 deaths). |
| **X Asymptomatic rate** | 0.02 (=2%) | Set at 2% (WHO 2020c) |
| **X Rate hospitalized patients (non-ICU)** | 0.484=<br>16,019 patients admitted to hospital (non-ICU)/33,089 total infected, at 22/03/2020) | Official data from the Spanish Ministry of Health (MSCBS 2020). |
| **X Rate ICU hospitalized patients** | 0.071= 2,355 ICU patients/33,089 total infected. | Official data from the Spanish Ministry of Health (MSCBS 2020). |

From the parameters (Tables 2 and 3) and the values in the *Stocks* (Table 1), we obtained the figures 3 to 7.

**Figure 3.**
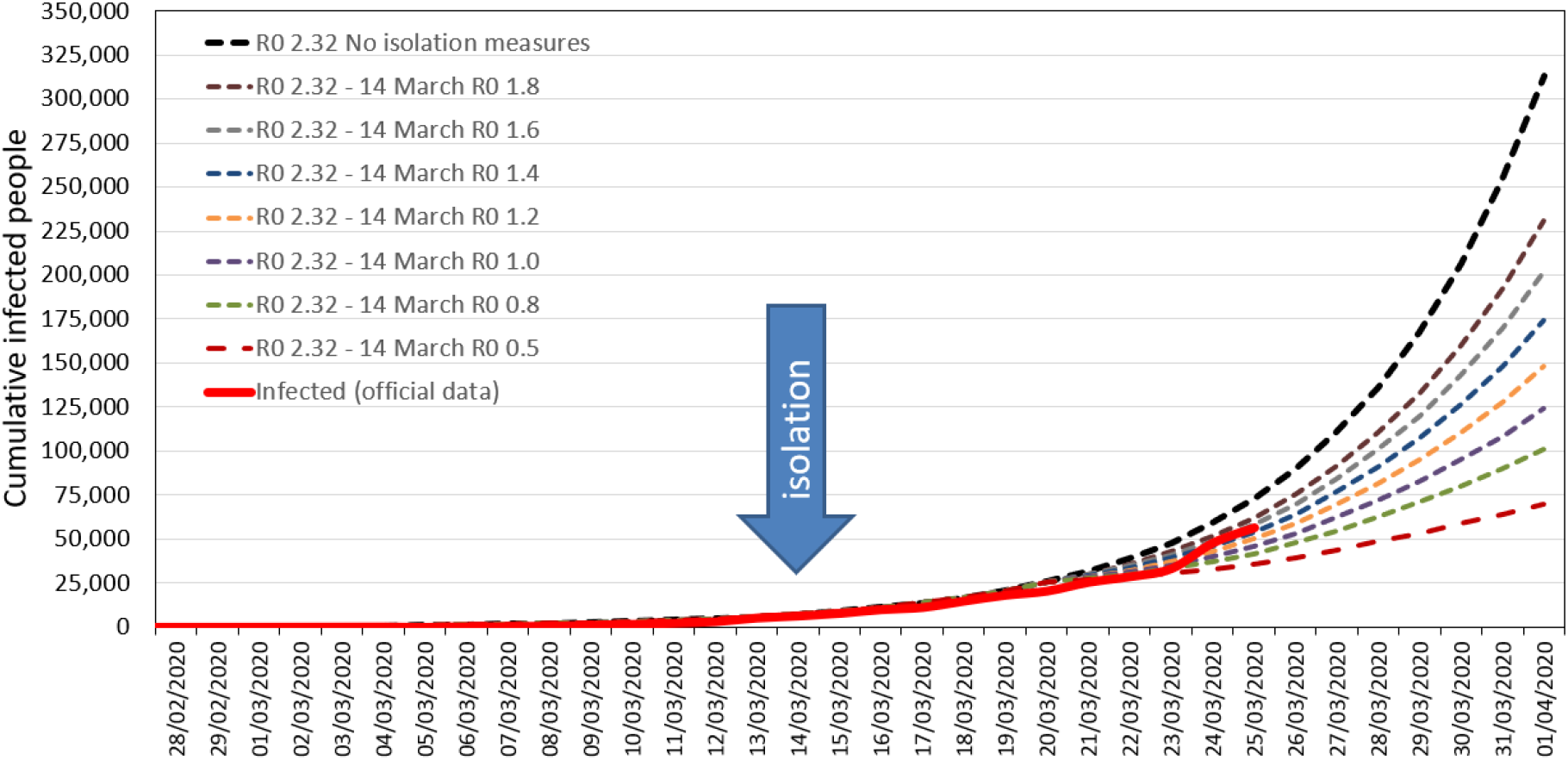
Confirmed cases of COVID-19 in Spain (red line) and data obtained from the model (dotted lines) with the parameters in Tables 1, 2 and 3. Isolation measures began on 14th March. See the effect of seven different R0 from 14^th^ March 2020.

Figure 3 shows the confirmed cases of COVID-19 in Spain (red line) and data obtained from the model (dotted lines) with the parameters in Tables 1, 2 and 3. Isolation measures began on 14th March. We added in this graph the effect of seven different R0 from the 14^th^ March and the expected evolution without any measures at all, maintaining a R0 of 2.32.

Figure 4 shows the estimated people infected (red dotted line) compared to the real data (red line) and the prediction of the model if R0 after 14^th^ March would be 0.8.

**Figure 4.**
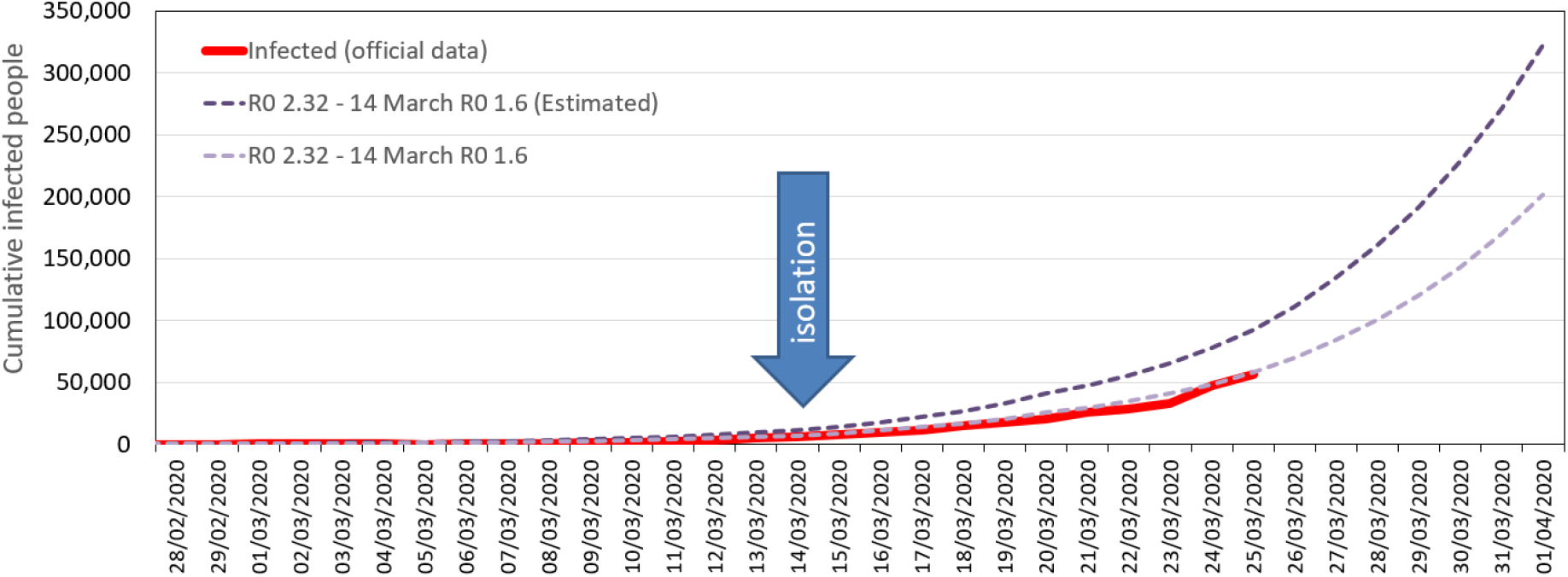
Infected people (official data, red line). Forecast of the model (violet dotted line) with a R0 of 2.32 until 13 March, and R0=1.6 from 14 March. The dark violet dotted line indicates the estimated real cases with a case fatality rate of 3.4%. The arrow points out the day that isolation measures were set by the Spanish Government (14 March 2020).

The adjustment of the number of deaths and fatality rate is shown in Figure 5. We had to apply a non-constant fatality rate to better fit the real numbers. Doted red and green lines show two scenarios, the first one with a higher and constant fatality rate, and the second, with a lower fatality rate after a peak.

**Figure 5.**
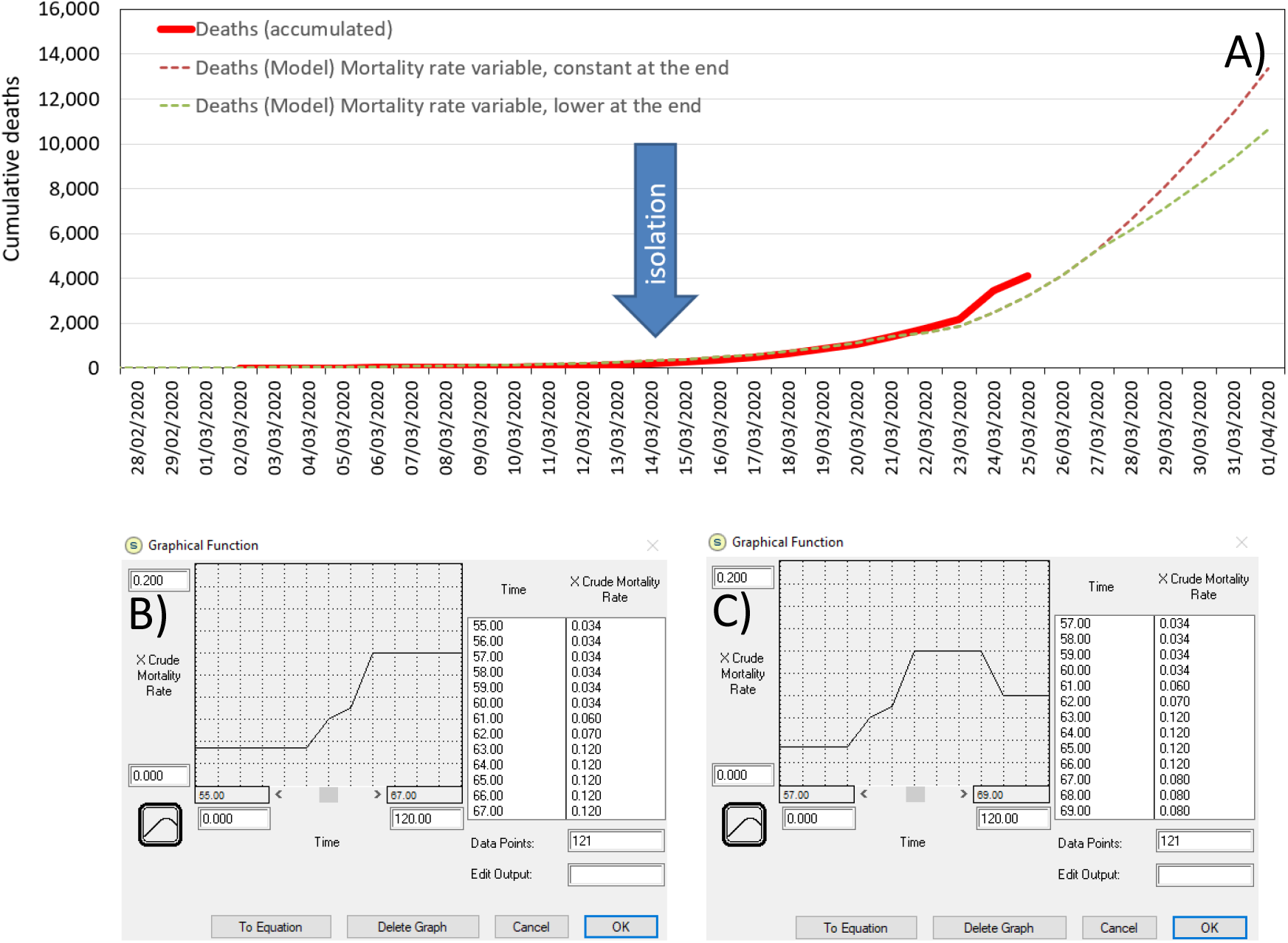
Calibration of the number of deaths. A) The red line shows the number of deaths up to 25 March. The death rate is not constant along the period. B) Table corresponds to the case fatality rate (oscillating between 3.4% and 12%) and death toll in the red dotted line in figure A. C) Table corresponds to the case fatality rate of the green dotted line in figure A.

## RESULTS

Once we calibrated the model, we built up different scenarios and compared them.

### Scenario 1. Implement isolation measures vs. no isolation at all

Isolation measures would affect the initial R0. In order to compare the effect of the implementation of different degrees of isolation measures, we set the same R0 at the beginning (until 14 March) and lower R0 after the lockdown.

A. Constant R0 of 2.32: no social distancing measures are implemented.
B. Initial R0 of 2.32, and R0 of 0 from 14 March, representing a theoretical scenario where there is not a single new infection after the lockdown.
C. Initial R0 of 2.32, and R0 of 0.1 from 14 March, representing hard social distancing measures.
D. Initial R0 of 2.32, and R0 of 0.8 from 14 March, representing medium social distancing measures.
E. Initial R0 of 2.32, and R0 of 1.5 from 14 March, representing milder social distancing measures.

### Scenario 2. Isolation measures one week after their actual adoption

We claim the usefulness of this approach to test theoretical scenarios. In Figure 7 we show the results of implementing the lockdown one week later (from 14 to 21 March). In order to get this scenario, we set the R0 from 21 March at 0.8.

### Scenario 3. Testing randomly all the population without symptoms on 14 March

From a basic scenario (R0 at 2.32 before 14 March and at 0.8 after that date), we analyzed the hypothetical action of randomly testing the asymptomatic people, i.e. uninfected and infected (asymptomatic and subclinical) people. In Figure 8, four scenarios are shown. All four refer to an unrealistic testing period of one day (14 March) when 5, 10, 20 and 30 million tests would be done. A positive test implies that a person goes into quarantine and hence, without any possibility to infect other people.

### Scenario 4. Ceasing or relaxing the isolation measures some days after the lockdown

From a basic scenario (R0 2.32 before 14 March and 0.8 from 14 March), we tested what 1) what would have happened if the isolation measures had ceased

2) what would have happened if the isolation measures had ceased or relaxed eight days after the lock down, that is 22 March, (R0 0.8 to 2.32) or get less severe (R0 0.8 to 1.4). Results are shown in Figure 9.

### Scenario 5. Predicting when would the peak of infections per day occur and the effect of different degrees of isolation measures

From the basic scenario (R0 2.32 before 14 March and 0.8 from 14 March), we predicted when the peak in infections per day would occur, and compared different R0 values, representing from harder isolation measures (lower R0) to more relaxed ones (higher R0). Figure 10 shows the result of the simulation.

## DISCUSSION

The entire world is facing a new pathogen, so far unknown for humans. Nevertheless, some experts had been lately suggesting that a similar scenario could happen (Allen et al. 2017).

Although the model has limitations, since it is a simplification of the reality, the results of the calibrated model (Figure 3) fits quite well with the official data of infected people with COVID-10 in Spain until 24 March. Until today, it is difficult to obtain the most probable R0 after 14 March because of the time lag of detected patients due to the incubation period (set at 6 days). Therefore, further time is needed to quantify the effect of mitigation measures in the R0 reduction.

We think that the real amount of infected people would reach, at least, 2.1 times the official numbers, calculated from a 3.4% fatality rate from days 24 and 25 March (Figure 4). If the fatality rate estimation changed, the real numbers of infected people would rise up consequently. It is important to remark that not knowing the real numbers of infected people does not significantly affect the power of the approach we propose here, since the comparison of different scenarios (e.g. reducing R0 after a day or another) uses the same base model. Hence, we can say that the measure that works better in the model, would work better in the real world as well. Besides, the results of this model would improve if we could adjust all the parameters to the real situation. Then, we could build a more robust model. In fact, our intention is to generate further versions of this model with whenever more accurate parameters are available.

Fatality numbers followed a smooth curve until 24 March, when the increase of deaths was higher than expected, but on 25 March it went down again. In case the amount of fatalities does not follow the way pointed out by the dotted lines in Figure 5A, we will recalculate the fatality rates showed in Figure 5A and 5B, but we will wait a few more days. Moreover, more time is still needed to check a possible third scenario, that is a higher case fatality rate as the epidemic saturates health services.

Regarding the social distancing measures, the model detects and quantifies the positive effect of reducing the R0, as seen in Figure 6. According to our results, the earlier the measures are taken, the better, as seen in Figure 7.

**Figure 6.**
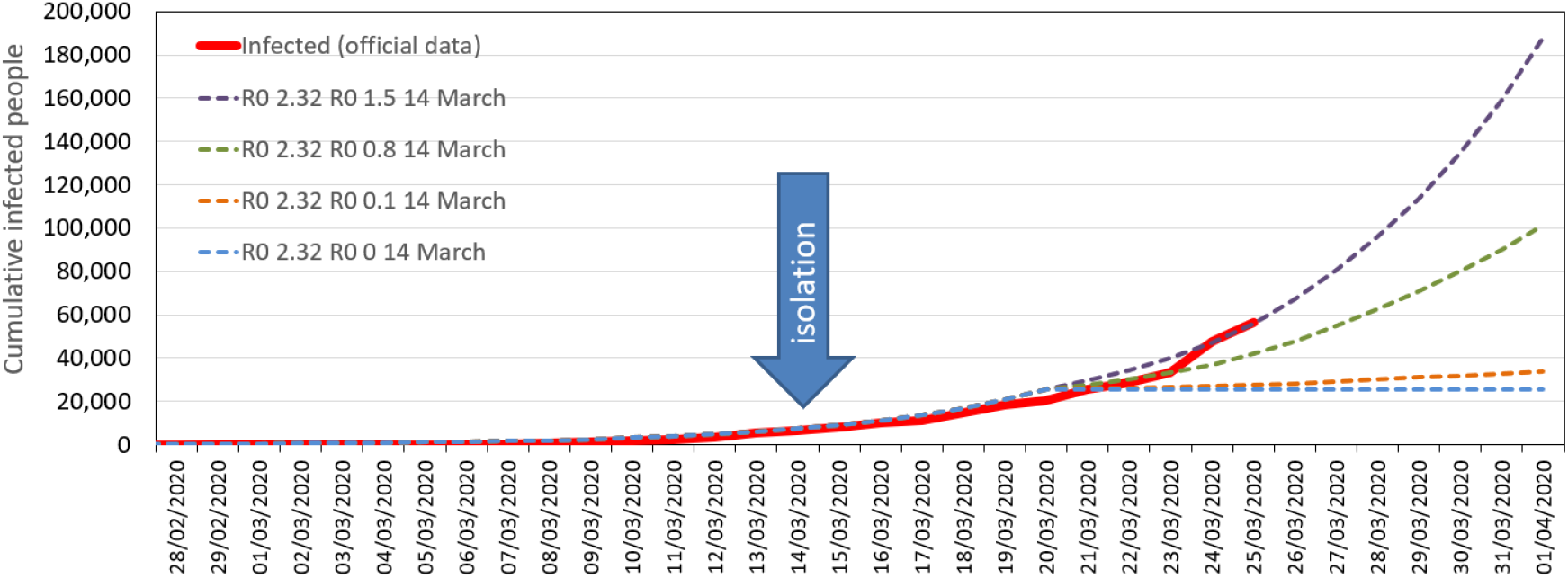
Scenario 1: Implemented isolation measures vs. no measures. Isolation measures would affect R0 from the day the measures are implemented. R0 of 2.32 during all the period represents no isolation measures. R0 0 and R0 0.1 represent hard isolation, being R0 0 the minimum theoretical, impossible to achieve. It seems that the R0 achieved after 14 March is around 1.5.

**Figure 7.**
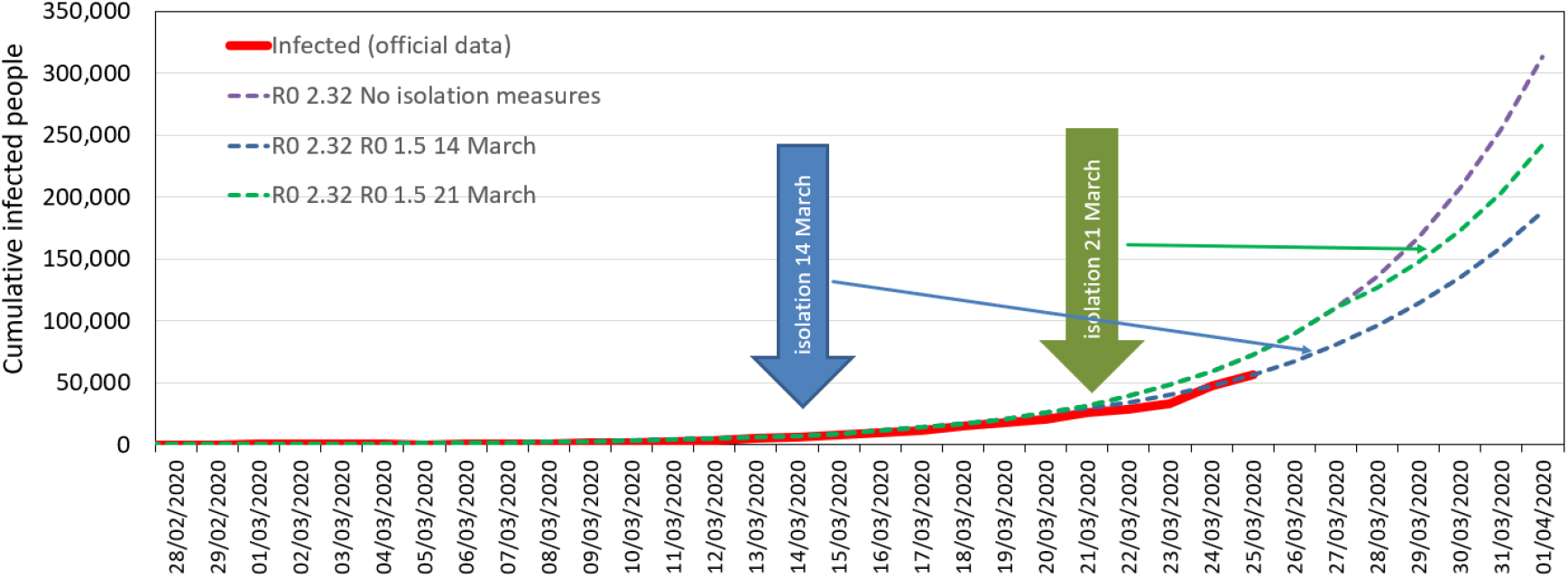
Scenario 2: Isolation measures implemented on 14 March vs. on 21 March using a R0 of 1.5 after the isolation measures’ onset. Violet dotted line shows no isolation measures taken.

We also tested what would happen if a random testing policy for all population would have been adopted. Even performing 20 or 30 million tests (from a population of 47 million) the result is poor (Figure 8), indicating that the efforts of testing should be more focused on suspected cases. This last approach was adopted in China by a contact tracing strategy (WHO 2020c) that allowed isolation of those infected prior to symptoms, and also a door-to-door monitoring to identify cases with symptoms (Shen et al. 2020).

**Figure 8.**
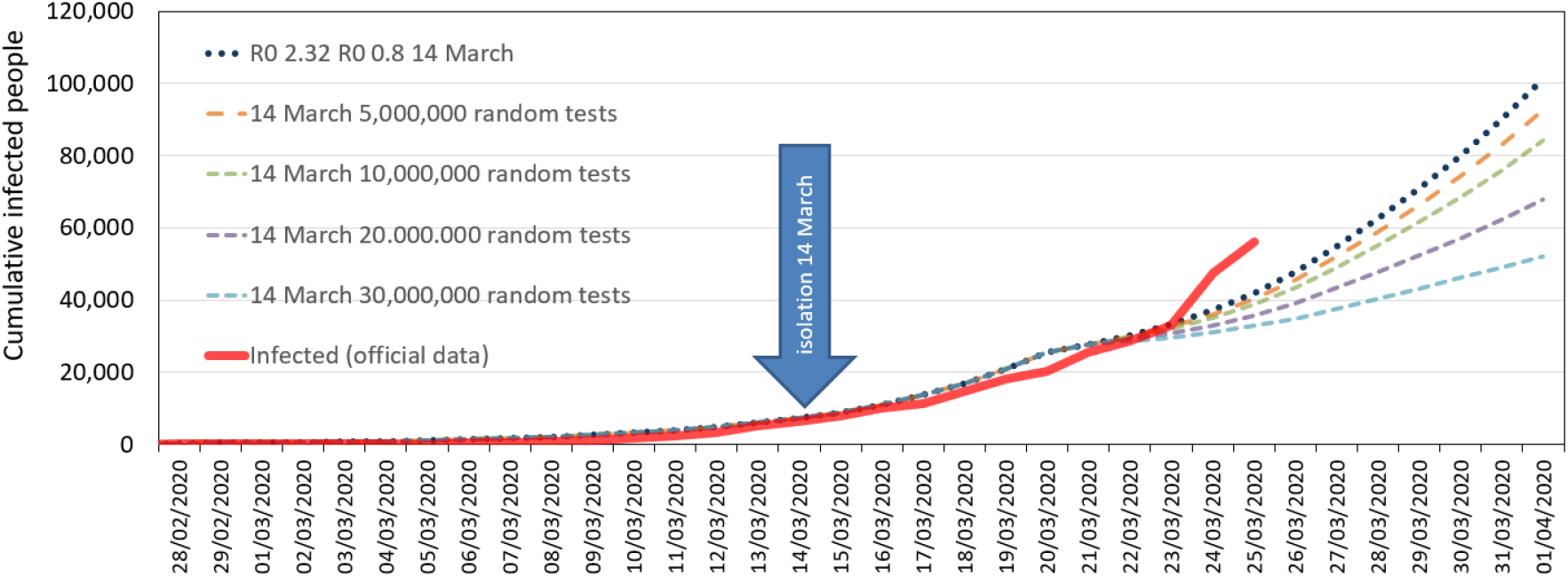
Scenario 3: Testing randomly people without symptoms and subclinical on 14 March. All scenarios with a R0 of 2.32 before 14 March, and of 0.8 afterwards. We hypothesized that if massive testing had been performed, R0 would have been kept at 0.8.

In the last of the scenarios, we analyzed what would happen if the isolation measures ceased 10 days after their implementation. As seen in Figure 9, these measures should not cease, since there are still infective individuals in the population and no herd immunization is operating due to the low percentage of population recovered and thus, immunized by illness.

**Figure 9:**
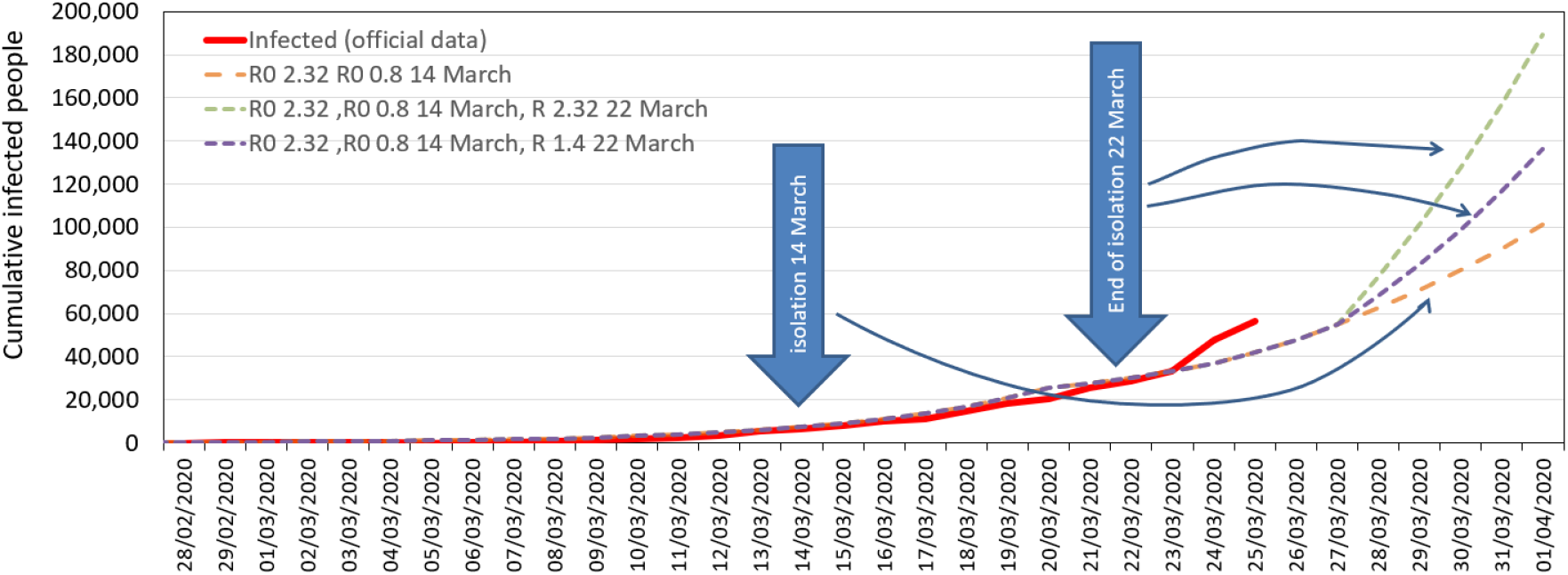
Scenario 4: Testing the effect of ceasing or relaxing the isolation measures. From a basic scenario (R0 2.32 before 14 March and 0.8 from 14 March), we tested what would happen if, 8 days after the lock down, the isolation measures ceased (R0 0.8 to 2.32) or turned into less severe measures (R0 0.8 to 1.4).

With the parameters used in the model, the peak of infected cases will occur around 8-9^th^ April (Figure 10). These numbers could be more accurate by using more reliable data of the COVID-19 evolution of the real infected population. Nonetheless, we think the error of the model could be of few days. Other models state that the peak would occur around 14^th^ April in Spain (Burgos-Simón et al. 2020).

**Figure 10:**
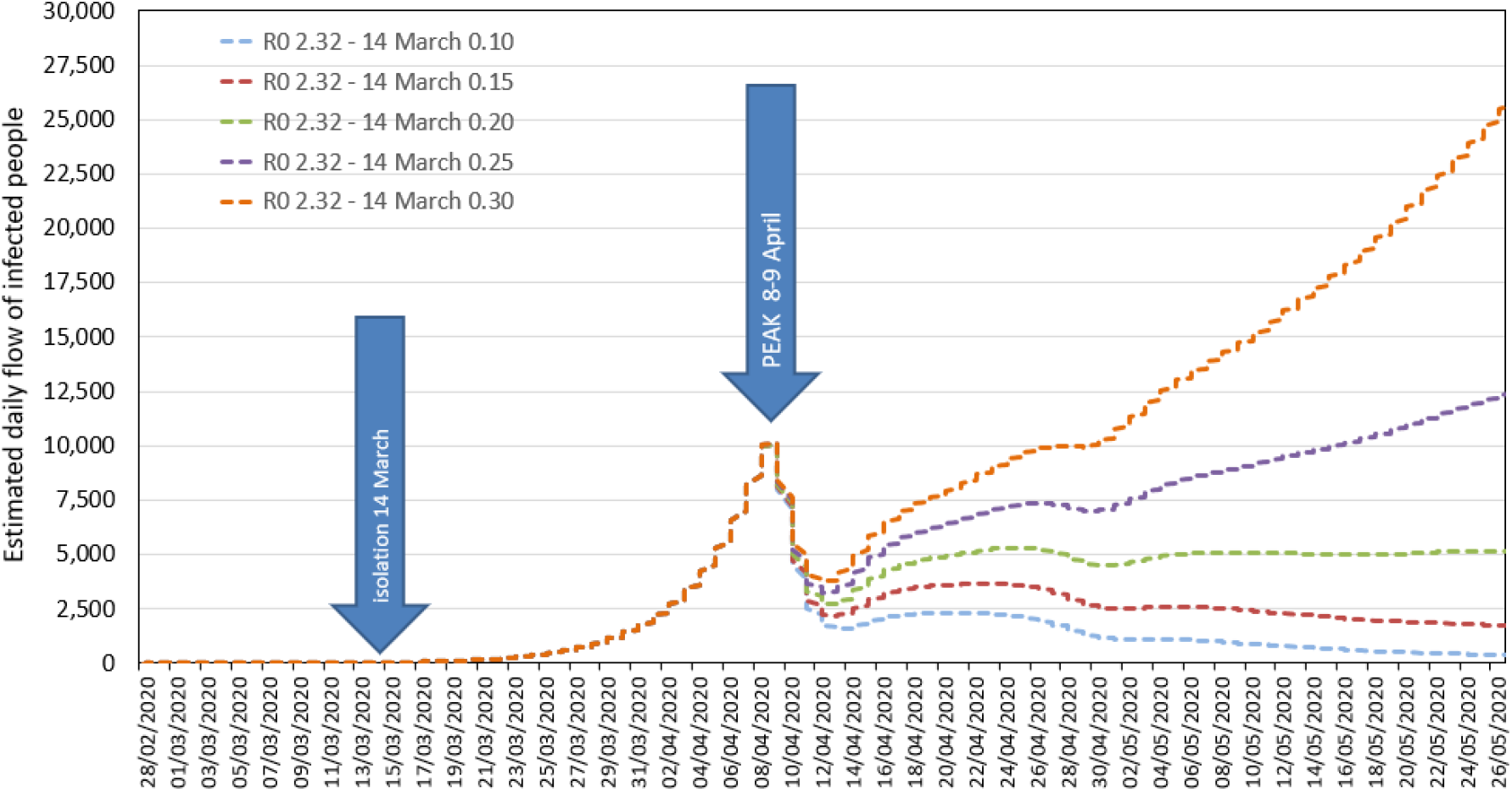
Scenario 5: Representation of estimated daily flow of infected people. Peak of infections per day is estimated to occur around 8-9 April. After that peak, it is crucial to maintain the isolation measures (lowering R0 as much as possible) in order to minimize the daily flow of new infections. A R0 below 0.15 would be desirable, i.e. a new case of infection every 6.7 infected cases.

It is interesting to point out that it would be vital to prevent infection number from increasing, maintaining the lowest R0 possible after the peak. As seen in Figure 10, an R0 higher than 0.15 would rise the number of new cases of infection again.

Other models of the spreading of COVID-19 in Spain, e.g. Burgos-Simón et al (2020), did not compare scenarios of different actions to reduce the speed of spreading and stated that the peak would occur between 18 and 20 April, about 10 days after our calculations (Figure 10). Arenas et al (2000) predicted ICU saturation from the end of March to the 23 April, although we did not represent the ICU patients, the peak of new infected people (Figure 10) is expected for the 8-9 April, so ICU higher demand will be concentrated from some weeks after that date, probably a couple of weeks after Arenas’ model. Another model applied in Spain with data from 28 February with few dozens of cases (Aleta and Moreno, 2020) expressed the necessity of early detection of positive cases with symptoms. But from our point of view, due to the incubation period when the patient is contagious and the great percentage of asymptomatic and subclinical cases, that strategy would have been not enough to stop the spread of this disease, now a pandemic.

Comparison of different scenarios has already been done by other researchers, such as Ferguson et al. (2020) for the United Kingdom, remarking that both models, ours and Ferguson’s, reflect the advantages of physical distance among people to reduce the number of infections.

This model does not pretend to impersonate official predictions nor scenarios about the COVID-19 spread in the different areas or countries. Considering the complexity of the spreading phenomenon and the exponential component, it is an impossible task for researchers and professionals interested in this pandemic to attempt to predict the dynamics or the effect of different measures without using a model that includes the main aspects of the pandemic. Regarding the forecast for Spain, there are further constraints on the model adjustment i.e. frequent changes in counting rules of infected people and case definition, and changes in testing and isolation policies. In fact, isolation measures were announced to become harder in Spain from Monday, 30 March.

Moreover, the evolution of the epidemic differs greatly from one Spanish region to another due to the spatial distribution of the people (rural areas vs. big cities), to population movements, and regional health systems’ characteristics, among others. It must be said that rates can be better calculated in samaller and more homogeneous areas and, in turn, better results can be obtained running the model.. Nevertheless, the model could be customized to fit any region following the instructions aforementioned.

One important limitation of our model is the lack of reliable data about the actual cases of infection, mainly asymptomatic and with mild symptoms. In the last days, only the severe cases and suspected cases among active health care staff have been confirmed with a test in Spain. We hope this situation can be changed in the following weeks, as the Spanish Government is acquiring more diagnostic tests (Spanish Government, 2020).

One strength of this model is that is has been conceived open-access. We keen to facilitate others to adapt the model to their own situation. The use of this model for an educational purpose can be a strong tool to convince people about the importance of adopting early social distancing approaches (Figure 7) and also to maintain them in time (Figure 8).

We think it could be particularly convenient for researchers and health care staff in low- and middle-income countries. These regions are especially vulnerable to the pandemic. Facing a new pathogen as CoV-2 and its dynamic represents a huge challenge for any country, especially for those with weaker health care systems. Some Sub-Saharan African countries have already detected more than 100 cases of COVID-19 (WHO 2020).

Although some knowledge about STELLA modeling is needed, we think that any interested person with a light training could customize it, by adapting the basic data (*Stocks* and *Converters*) to the region of interest and generate different scenarios. We are now working in a video tutorial to explain the design of the model in order to facilitate other stakeholders to modify the structure and customize parameters to their areas of interest.

***

This version was finished 27 March.

### New versions

Taking into consideration the rapid spread of the virus and the changing numbers, we intent to publish new versions of the model every few days, which would include improvements in structure and modeling of new scenarios.

### Collaboration and training

Other contributing authors interested in this COVID-19 spreading model are welcomed. Our intention is to improve the model both in the structure and parameters to make it more accurate and useful.

Researchers with a substantial and sustained collaboration are welcomed as co-authors. We are also opened to train and help other users to use this model (video tutorial coming soon).

## Data Availability

All data are completely available to other researchers

## Acknowledgements

We wish to thank Marta Bordehore for her valuable comments.

## Contributions

C.B.: model concept, STELLA® programing, write the manuscript, approved the submitted version.

M.N., Z.H.: model concept, data acquisition, write the manuscript, approved the submitted version.

E.S.F: write the manuscript, data acquisition, approved the submitted version.

## Notes

### Competing Interest Statement

The authors have declared no competing interest.

### Funding Statement

Research funded by the University of Alicante, Spain

